# Unmasking selective path integration deficits in Alzheimer’s disease risk carriers

**DOI:** 10.1101/19009662

**Authors:** Anne Bierbrauer, Lukas Kunz, Carlos A. Gomes, Maike Luhmann, Lorena Deuker, Stephan Getzmann, Edmund Wascher, Patrick D. Gajewski, Jan G. Hengstler, Marina Fernandez-Alvarez, Mercedes Atienza, Davide M. Cammisuli, Francesco Bonatti, Carlo Pruneti, Antonio Percesepe, Youssef Bellaali, Bernard Hanseeuw, Bryan A. Strange, Jose L. Cantero, Nikolai Axmacher

## Abstract

Alzheimer’s disease (AD) manifests with progressive memory loss and spatial disorientation. Neuropathological studies suggest early AD pathology in the entorhinal cortex (EC) of young adults at genetic risk for AD (*APOE* ε4-carriers). Because the EC harbors grid cells, a likely neural substrate of path integration (PI), we examined PI performance in *APOE* ε4-carriers during a virtual navigation task. We report a selective impairment in *APOE* ε4-carriers specifically when recruitment of compensatory navigational strategies via supportive spatial cues was disabled. A separate fMRI study revealed that PI performance was associated with the strength of entorhinal grid-like representations, suggesting grid cell dysfunction as a mechanistic explanation for PI deficits in *APOE* ε4-carriers. Furthermore, retrosplenial cortex was involved in the recruitment of compensatory navigational strategies via supportive spatial cues. Our results provide evidence for selective PI deficits in AD risk carriers, decades before potential disease onset.

## Introduction

Alzheimer’s disease (AD), by far the most common form of dementia (Querfurth and LaFerla, 2010), is characterized by a progressive deterioration of cognitive functions, starting with episodic memory loss and spatial disorientation (Kunz *et al.*, 2015; Coughlan *et al.*, 2018). No causal therapies for AD are currently available, possibly because drugs that would otherwise be effective are applied too late (Sperling, Clifford and Aisen, 2011). Hence, developing biomarkers and behavioral tests for identifying subjects at risk for developing AD is a crucial goal of current AD research.

The ε4 allele of the apolipoprotein E gene (*APOE*) is the most important genetic risk factor for late-onset AD (Corder *et al.*, 1993). Thus, it may provide an opportunity for assessing subclinical alterations of behavior, brain structure and function at very early disease stages (Matura *et al.*, 2014; Coughlan *et al.*, 2018). However, prior studies and meta-analyses on *APOE*-behavior relationships in young and middle-aged healthy participants showed divergent results: While some described cognitive impairments (e.g., Rawle *et al.*, 2018; Coughlan *et al.*, 2019), others reported improved functioning (suggesting antagonistic pleiotropy; e.g., Stening *et al.*, 2016; Zokaei *et al.*, 2017), or no effects (e.g., Matura *et al.*, 2014; Weissberger *et al.*, 2018; Coughlan *et al.*, 2019).

Here, we hypothesized that these divergent findings occur because *APOE* ε4 effects on behavior are mediated AD pathology in confined brain regions, which may be compensated by increased recruitment of unaffected areas - possibly resulting in altered cognitive strategies. Indeed, postmortem brain studies revealed first signs of neurodegeneration in the form of neurofibrillary tangles in *APOE* ε4-carriers already in early adulthood (Ghebremedhin *et al.*, 1998). Because tau-related neurodegeneration correlates closely with cognitive dysfunction (Giannakopoulos *et al.*, 2000; Jagust, 2018; Hanseeuw *et al.*, 2019), such very early tauopathy may manifest in subtle behavioral alterations.

Among the first regions to be affected by neurofibrillary tangles is the entorhinal cortex (EC) (Braak *et al.*, 2011), a hub for spatial navigation and memory. The EC contains spatially modulated cell types, including grid cells that fire at the vertices of equilateral triangles tiling the environment (Hafting *et al.*, 2005). In AD mouse models, early tauopathy in EC impairs grid cell functioning and spatial memory performance (Fu *et al.*, 2017). In humans, grid cell activity can be indirectly measured as “grid-like representations” (GLRs) via functional magnetic resonance imaging (fMRI) (Doeller, Barry and Burgess, 2010; Bellmund *et al.*, 2016; Kunz *et al.*, 2019). These GLRs were described to be functionally relevant for memory and spatial navigation (Doeller, Barry and Burgess, 2010; Kunz *et al.*, 2015; Stangl *et al.*, 2018). We previously showed evidence for impaired GLRs in young *APOE* ε4-carriers during virtual navigation in an arena (Kunz *et al.*, 2015). The spatial navigation performance of risk carriers was preserved, suggesting the employment of compensatory strategies. Indeed, reduced GLRs were accompanied by relatively increased hemodynamic activity in the hippocampus (HC). Moreover, *APOE* ε4-carriers navigated more often at the border of the arena, possibly in an attempt to stabilize their GLRs (Hardcastle, Ganguli and Giocomo, 2015; Kunz *et al.*, 2015; Coughlan *et al.*, 2019).

Together, these findings suggest that subtle alterations in the neural and behavioral signatures of spatial navigation may occur in *APOE* ε4-carriers already at an early age, and such changes may constitute prime candidates for neurocognitive markers of AD (Coughlan *et al.*, 2018). However, in order to dissect navigational strategies, potential compensatory mechanisms, and their underlying neural computations, it is crucial to probe the navigational function of grid cells more directly.

Theoretical considerations (Hafting *et al.*, 2005; Bush *et al.*, 2015), computer models (Burak and Fiete, 2009; Banino *et al.*, 2018), and empirical studies in rodents (Gil *et al.*, 2018) and humans (Stangl *et al.*, 2018) suggest that grid cells particularly support path integration (PI) processes. PI is the process of estimating one’s current position based on information about previous positions, heading direction, speed, and time elapsed (Gallistel, 1990). Specifically, grid cells may provide the computational basis for representing the integrated path (Burak and Fiete, 2009) and for computing direct vectors to spatial goals (O’Keefe and Burgess, 2005; Kubie and Fenton, 2012; Howard *et al.*, 2014; Stemmler, Mathis and Herz, 2015; Epstein *et al.*, 2017). Accurate goal vectors are essential for the “incoming phase” of PI tasks in which subjects have to return to their home location.

When only information from visual flow is available, PI inevitably leads to error accumulation (Gallistel, 1990; Burak and Fiete, 2009; Hardcastle, Ganguli and Giocomo, 2015). This suggests that sparse environments and longer navigational routes unmask subtle PI deficits in *APOE* ε4-carriers. By contrast, when supportive spatial cues such as boundaries or landmarks are available (as in most previous studies: Kunz *et al.*, 2015; Coughlan *et al.*, 2019), additional brain regions could be recruited, in particular HC (Doeller, King and Burgess, 2008; Kunz *et al.*, 2015) and retrosplenial cortex (RSC) (Mitchell *et al.*, 2018). This may enable *APOE* ε4-carriers to perform on par with control participants.

In line with these predictions, our study unmasks a specific deficit of *APOE* ε4-carriers in PI when no supportive spatial cues are available, and identifies the neural mechanisms underlying this deficit.

## Results

### Experimental task and study sample

We assessed PI performance with a novel task, the “Apple Game” (**Figures 1** and **S1**; Methods; see also **Video S1**). In each trial, participants first navigated to a basket (“start phase”), whose location they were instructed to remember (“goal location”). Next, participants navigated towards a variable number of trees (“outgoing phase”), systematically varying outgoing distance (and thereby PI difficulty), until they found a tree with an apple (“retrieval location”). Basket and trees appeared consecutively and disappeared as soon as the participant reached their locations. From the retrieval location, participants had to take the shortest route back to the basket location (“incoming phase”). Importantly, in different subtasks, participants either had to rely purely on visual flow (“pure PI”, PPI) or were provided with supportive spatial cues in the form of a boundary (“boundary-supported PI”, BPI) or an intramaze landmark (“landmark-supported PI”, LPI). PI performance was quantified via the distance between response location and correct location of the basket (“drop error”; for alternative metrics, see **Figure 1D**).

**Figure 1.**
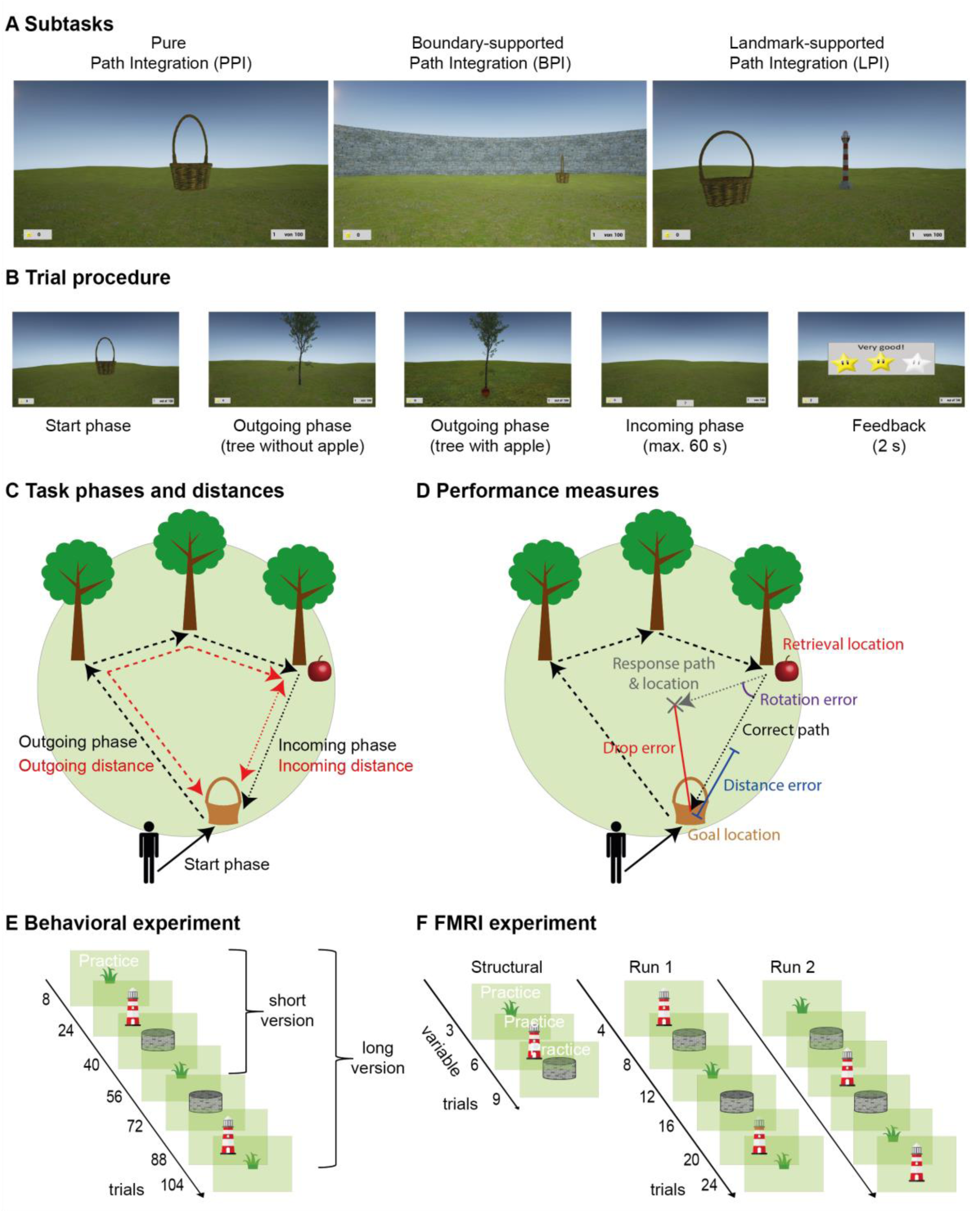
Experimental paradigm. (**A**) Participants performed a novel PI task (the “Apple Game”) in a virtual environment. The task comprised three subtasks that differed with regard to the presence or absence of supportive spatial cues: the pure PI (PPI) subtask without any supportive cue; the boundary-supported PI (BPI) subtask with a circular boundary; and the landmark-supported PI (LPI) subtask with an intra-maze landmark (lighthouse) close to the center of the environment. (**B**) In each trial, participants collected a basket (start phase) and tried to remember its location (goal location). After navigating towards a variable number of trees (1-5; outgoing phase), which disappeared after having been reached, participants had to find back to the goal location (incoming phase). Finally, they received feedback via different numbers of stars, depending on response accuracy. (**C**) Outgoing distance refers to the cumulated distance during the outgoing phase, incoming distance refers to the Euclidean distance between retrieval location (tree with apple) and goal location (basket). (**D**) PI performance was assessed as the distance between the correct goal location and the response location (drop error). The drop error can be separated into the distance error (i.e., the difference between the retrieval-to-goal distance and the retrieval-to-response distance) and the rotation error (i.e., the difference between the retrieval-to-goal rotation and the retrieval-to-response rotation). (**E**) The behavioral task comprised 8 practice trials followed by 16 trials in each subtask (short version; in the long version, all subtasks were performed twice, resulting in 32 trials in each subtask). (**F**) The fMRI task consisted of up to 9 practice trials during the structural scan, followed by two functional runs with six blocks of four trials each. See also **Figure S1, Video S1**.

We conducted the paradigm in *N*=267 healthy participants genotyped for *APOE* across four different European sites (“*APOE* sample”; *n*=65 *APOE* ε3/ε4 carriers, risk group; *n*=202 *APOE* ε3/ε3 carriers, control group; age range, 18-75 years; mean age, 37.7 years; 38.6% male). Control and risk group did not differ regarding demographic characteristics (**Table S2, Figure S2**). For a subgroup of this sample, structural MRI data were available, allowing us to establish relationships between behavioral performance and brain structure (“sMRI sample”; *n*=99 participants; *n*=23 risk carriers, *n*=76 controls). Furthermore, we recruited a separate group of *n*=35 participants who completed a variant of the task inside the MR scanner (“fMRI sample”). For sample details, see **Tables S1-S3**.

### Determinants of PI performance

We analyzed PI performance as a function of *APOE* genotype, subtask, and path distance in the *APOE* sample. We used a series of linear mixed models with “subtask” (PPI, BPI, or LPI) and “path distance” as within-subject variables and “*APOE*” as between-subjects variable. Path distance refers to either “outgoing distance” (i.e., the accumulated path distance during the outgoing phase; **Figure 1C**; Model 1a; **Table 1**; Methods) or “incoming distance” (i.e., the Euclidean distance between retrieval location and goal location; **Figure 1C**; Model 1b). When reporting inference statistics, “all” or “both” always refer to the two models with either one of the two path distances. “Subject” and “site” were added as random factors, and “sex” and “age” as covariates (for main effects and for interactions with subtask and genotype). Post-hoc comparisons were Tukey-corrected for multiple comparisons (number of subtasks). We found main effects of subtask and both path distance measures on PI performance (all *F* ≥ 466.42, all *P* < 0.001 for the two models with incoming and outgoing distance; **Figure S3**). Pairwise comparisons showed that performance was worse in the PPI as compared to the BPI and the LPI subtasks (all *z* ≥ 22.25, all *P*_Tukey_ < 0.001). Performance in the BPI subtask was also worse than in the LPI subtask (both *z* ≥ 6.44, both *P*_Tukey_ < 0.001).

**Table 1.** Statistical models.

| Model | Criterion | Predictors<br>(within subject) | Predictors<br>(between subject) | Covariates | Random<br>effects |
| --- | --- | --- | --- | --- | --- |
| **Effects of *APOE*, subtask, and path distance on performance** |  |  |  |  |  |
| **1a** | Performance | Subtask,<br>Outgoing distance | *APOE* | Age, Sex | Subject,<br>Site |
| **1b** | Performance | Subtask,<br>Incoming distance | *APOE* | Age, Sex | Subject,<br>Site |
| **1c** | Performance | Subtask,<br>(Outgoing and<br>incoming)<br>path distance,<br>Type of distance | *APOE* | Age, Sex | Subject,<br>Site |
| **Effects of landmark and boundary** |  |  |  |  |  |
| **2a** | Performance | Goal-to-boundary<br>distance | *APOE* |  | Subject,<br>Site |
| **2b** | Performance | Goal-to-landmark<br>distance | *APOE* |  | Subject,<br>Site |
| **2c** | Distance to<br>boundary |  | *APOE* |  | Subject,<br>Site |
| **2d** | Distance to<br>landmark |  | *APOE* |  | Subject,<br>Site |
| **Effects of EC volume, *APOE*, subtask, and path distance on performance** |  |  |  |  |  |
| **3a** | Performance | Subtask,<br>Outgoing distance | *APOE*,<br>Relative volume in EC | Age, Sex | Subject |
| **3b** | Performance | Subtask,<br>Incoming distance | *APOE*,<br>Relative volume in EC | Age, Sex | Subject |
| **Effects of HC volume, *APOE*, subtask, and path distance on performance** |  |  |  |  |  |
| **4a** | Performance | Subtask,<br>Outgoing distance | *APOE*,<br>Relative volume in HC | Age, Sex | Subject |
| **4b** | Performance | Subtask,<br>Incoming distance | *APOE*,<br>Relative volume in HC | Age, Sex | Subject |
| **Effects of RSC volume, *APOE*, subtask, and distance on performance** |  |  |  |  |  |
| **5a** | Performance | Subtask,<br>Outgoing distance | *APOE*,<br>Relative volume in RSC | Age, Sex | Subject |
| **5b** | Performance | Subtask,<br>Incoming distance | *APOE*,<br>Relative volume in RSC | Age, Sex | Subject |
| **Mechanistic model to explain path integration performance** |  |  |  |  |  |
| **6** | Performance | Subtask,<br>Incoming distance | EC integrated path<br>representations during<br>incoming phase,<br>HC goal proximity<br>representations during<br>incoming phase,<br>pmEC GLRs,<br>RSC landmark<br>representations |  | Subject |

Notably, incoming distance had a more pronounced effect in the PPI subtask than in the two other subtasks, as shown by a significant subtask by incoming distance interaction (*F* = 99.17, *P* < 0.001, **Figure S3**). This was not the case for outgoing distance (*F* = 1.02, *P* = 0.361). Post-hoc analyses confirmed that incoming distance had a stronger effect on performance in the PPI subtask than in the two other subtasks (both *z* ≥ 9.80, both *P*_Tukey_ < 0.001), with no difference between BPI and LPI (*z* = 1.11, *P*_Tukey_ = 0.511). Furthermore, we encountered significant main effects of sex and age (all *F* ≥ 63.49, all *P* < 0.001): Younger age and male sex predicted better performance [see **Figure S4** (also for interactions involving covariates)].

These results show that the absence of supportive spatial cues not only reduces PI performance, but also increases the impact of longer incoming distances, which is presumably related to larger error accumulation (Gallistel, 1990; Burak and Fiete, 2009; Hardcastle, Ganguli and Giocomo, 2015).

### *APOE* effects on PI are unmasked in the absence of supportive spatial cues

We next analyzed effects of *APOE* on performance. No main (subtask-independent) effect of *APOE* was observed (both *F* ≤ 0.04, both *P* ≥ 0.850), in line with a previous study showing generally unimpaired PI performance in *APOE* ε4-carriers^10^. Importantly, however, we observed a significant *APOE* by subtask interaction (both *F* ≥ 12.60, both *P* < 0.001; **Figure 2A**), indicating different *APOE* effects on performance in the individual subtasks. The interaction was driven by worse performance of risk carriers as compared to controls specifically in the PPI subtask (both *z* ≥ 2.44, both *P*_Tukey_ ≤ 0.039), while there was no difference in the BPI (both *z* ≤ 0.05, both *P*_Tukey_ ≥ 0.998) or the LPI subtasks (both *z* ≤ 0.98, both *P*_Tukey_ ≥ 0.593). To scrutinize this result, we compared performance as a function of *APOE* between the PPI subtask and either the BPI or the LPI subtask, respectively (**Figure 2B**). These contrasts measure a specific impairment of PI performance in the absence of supportive spatial cues. Indeed, the performance of risk carriers declined more strongly than that of controls in the PPI subtask when compared to either the BPI subtask (both *z* ≥ 2.86, both *P*_Tukey_ ≤ 0.012) or the LPI subtask (both *z* ≥ 4.10, both *P*_Tukey_ < 0.001). Post-hoc analyses revealed that the effect of genotype on performance was driven by increased rotation errors (**Figure 1D**; **Fig. S5**).

**Figure 2.**
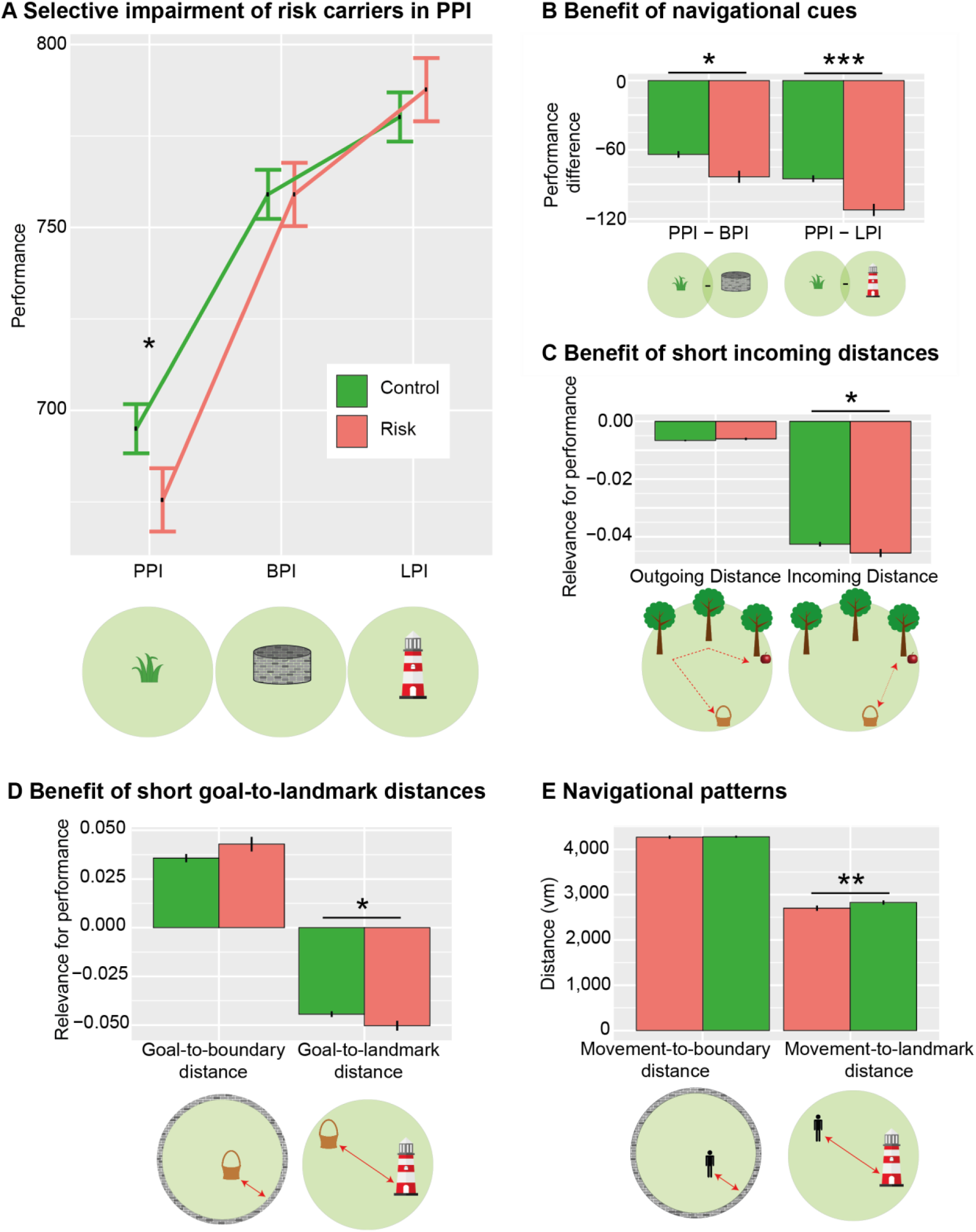
Performance as a function of genotype, distances, and subtask. (**A**) Performance (which is inversely related to drop error) is specifically impaired in risk carriers when no supportive spatial cues are available, i.e., in the PPI subtask. (**B**) Risk carriers benefit more from environmental landmarks and boundaries than controls. (**A, B**) depict results from model 1b; results from model 1a are statistically equivalent. (**c**) Incoming (but not outgoing) distance is more closely related to spatial memory performance in risk carriers than controls (model 1c). (**D**) Goal-to-landmark distance (but not goal-to-boundary distance) is more relevant in risk carriers than controls (models 2a, b). (**E**) Movement-to-landmark distance (but not movement-to-boundary distance) is significantly lower in risk carriers than controls (models 2c, d). Y-axes show parameter estimates resulting from the different models; error bars, s.e.m.; \**P* < 0.05; \*\**P* < 0.01; \*\*\**P* < 0.001; Control, *APOE* ε3/ε3-carriers; Risk, *APOE* ε3/ε4-carriers; PI, path integration, PPI, pure path integration; BPI, boundary-supported path integration; LPI, landmark-supported path integration; vm, virtual meters. See also **Figures S2, S3, S4, S5, Tables S1, S2, S5**.

Moreover, we observed an interaction between *APOE* and incoming distance (*F* = 3.94, *P* = 0.047; **Figure 2C**), presumably reflecting stronger error accumulation (Gallistel, 1990; Burak and Fiete, 2009; Hardcastle, Ganguli and Giocomo, 2015) in risk carriers than controls. This effect was not significant for outgoing distance (*F* = 1.86, *P* = 0.173). Indeed, *APOE* effects were significantly stronger for incoming than outgoing distance (three-way interaction between outgoing distance, incoming distance, and *APOE* in a model including both distance types; *F* = 4.68, *P* = 0.030; Model 1c). This indicates that performance in risk carriers is particularly affected by the incoming (i.e., Euclidean) distance between retrieval and goal location.

Since risk carriers’ performance was on par with controls when supportive spatial cues were present, we hypothesized that the distance to these cues determines their performance more than in controls. We thus investigated PI performance as a function of “spatial cue distance” (i.e., the distance between the goal location and the boundary or the landmark, respectively; Models 2a,b). Generally, higher distances from the boundary and lower distances from the landmark improved performance (both *F* > 342.45, both *P* < 0.001). Additionally, the performance of risk carriers declined more strongly with increasing goal-to-landmark distance than the performance of controls (*F* = 4.55, *P* = 0.033, Model 2b; **Figure 2D**). By contrast, goal-to-boundary distance did not exhibit different effects in risk and control participants (*F* = 2.59, *P* = 0.107; Model 2a; **Figure 2D**).

Irrespective of PI performance, we investigated the mean distance of participants’ navigation to the boundary or the landmark, respectively (only during the incoming phase, as movements during the outgoing phase were determined by the trees; Models 2c,d). This revealed that risk carriers showed a different navigational pattern: They navigated in closer proximity to the landmark (*F* = 6.98, *P* = 0.009; Model 2d, **Figure 2E**), suggesting that they used the landmark information to a higher degree to guide their behavior. By contrast, risk carriers navigated at similar distances to the boundary as controls (*F* = 0.06, *P* = 0.803; Model 2c, **Figure 2E**). In sum, a selective PI deficit in *APOE* ε4-carriers was unmasked when environments lacked spatial cues and participants had to rely purely on PI. When environmental landmarks were available, risk carriers relied more strongly on their proximity.

### Brain structural determinants of *APOE* effects on PI

How is performance in the different subtasks related to brain volume in areas relevant for spatial navigation, and how are these relationships modulated by *APOE*? We analyzed PI performance as a function of relative gray matter volume in three regions of interest (ROIs): EC, HC, and RSC. We used mixed models with “subtask” and “path distance” (“outgoing distance”, Models 3a-5a; or “incoming distance”, Models 3b-5b) as within-subject variables; “*APOE*” and “volume” of one of the three ROIs as between-subject variables; and “subject” as random factor (sMRI sample; **Table 1**; Methods). “Age” and “sex” were added as covariates. Because these models contain two continuous predictors of interest (path distance and volume), post-hoc comparisons were performed on quintiles of the path distance predictor (Tukey-corrected for number of compared quintiles). Qualitatively identical results were obtained when the path distance predictor was subdivided into quartiles or tertiles (**Supplementary Results**).

Risk carriers and controls did not differ regarding age, sex, whole brain volume, and relative volume in any ROI (**Table S3, Figure S2**), in line with previous findings (Matura *et al.*, 2014). Also, relative ROI volumes did not directly predict performance in any subtask, for either controls or risk carriers (all *z* ≤ 1.68, all *P* ≥ 0.093). Importantly, however, an effect of EC volume on performance in risk carriers was unmasked when analyzed in relationship to incoming distance: When we examined the effect of EC volume on performance (Model 3b), we encountered a significant interaction with *APOE* and incoming distance (*F* = 6.70, *P* = 0.010, **Figure 3**). While EC volumes did not predict performance for shorter incoming distances in either risk carriers or controls [800 virtual meters (vm)–2580 vm, all *z* ≤ 1.24, all *P*_Tukey_ ≥ 0.214], they exerted a significant effect for longer incoming distances specifically in risk carriers (4360 vm–7920 vm; all *z* ≥ 2.17, all *P*_Tukey_ ≤ 0.030), but not in controls (all *z* ≤ 1.87, all *P*_Tukey_ ≥ 0.061). Indeed, effects in risk carriers were significantly larger than in controls at large incoming distances (7920 vm; *z* = 2.56, *P*_Tukey_ = 0.044), but not at lower incoming distances (800 vm-6140 vm, all *z* ≤ 2.13, all *P*_Tukey_ ≥ 0.126). No comparable effects were found for outgoing distance (Model 3a), in HC (Model 4a/4b), or RSC (Model 5a/5b). These results show a selective influence of EC volume on performance in risk carriers for longer incoming distances, suggesting that effects of subtle structural degradation of the EC in risk carriers become visible in situations of large potential error accumulation.

**Figure 3.**
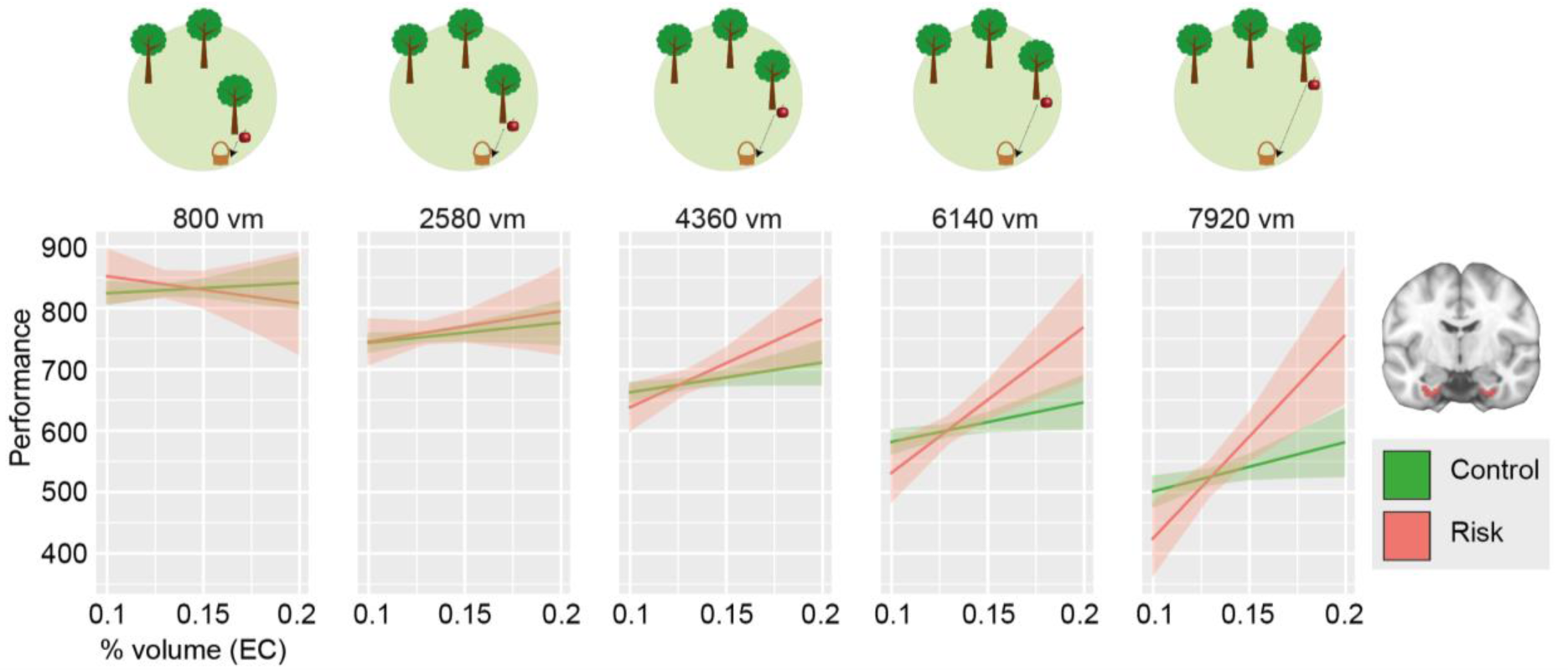
Performance as a function of relative EC gray matter volume, incoming distance and genotype. EC gray matter volume predicts performance only during PI with long incoming distances (middle to right panels) and only in risk carriers (model 3b). As the model contained two continuous predictors, one of them (incoming distance) was discretized into quintiles for post-hoc tests and for graphical depiction. Y-axis shows parameter estimates for performance; shaded area, s.e.m.; control, *APOE* ε3/ε3-carriers; risk, *APOE* ε3/ε4-carriers; PI, path integration; % volume, percent of whole brain volume; vm, virtual meters. See also **Table S3**.

### FMRI correlates of integrated path and goal proximity during PI

In order to understand the neural basis of PI and the impact of supportive spatial cues in our paradigm, we conducted an fMRI study with an adapted version of the task (fMRI sample; Methods). First, we analyzed neural representations of two fundamental metrics of PI: the instantaneous integrated path (i.e., the length of the travelled path during either the outgoing or the incoming phase) and the instantaneous goal proximity (i.e., the distance between the subject’s current location and the goal). We created a general linear model (GLM) with regressors for the entire time of movement during the outgoing and the incoming phase, separately for each subtask (**Figure S6**). In two separate analyses, we included either integrated path or goal proximity as time-varying parametric modulators during movement (sampled at a temporal resolution of 5Hz). We tested how these measures of PI were related to BOLD activity in the three ROIs (EC, HC, RSC). *P*-values were FDR-corrected for multiple comparisons (number of ROIs and number of contrasts).

During the outgoing phase, EC and HC showed pronounced deactivation with increasing integrated path (both *t*_34_ ≥ 4.68, both *P*_FDR_ < 0.001; **Figure 4A**). This was reversed during the incoming phase, where we observed increased activation in both areas with integrated path (both *t*_34_ ≥ 2.55, both *P*_FDR_ ≤ 0.046; **Figure 4A**). When analyzed separately for the different subtasks, we found that EC and HC activity levels always differed significantly from zero in the PPI subtask (**Supplementary Results**; **Figure 4C**). By contrast, integrated path did not predict activity in the LPI subtask, suggesting that this representation is not relevant in this subtask. Activity in RSC did not show any relationship with integrated path.

**Figure 4.**
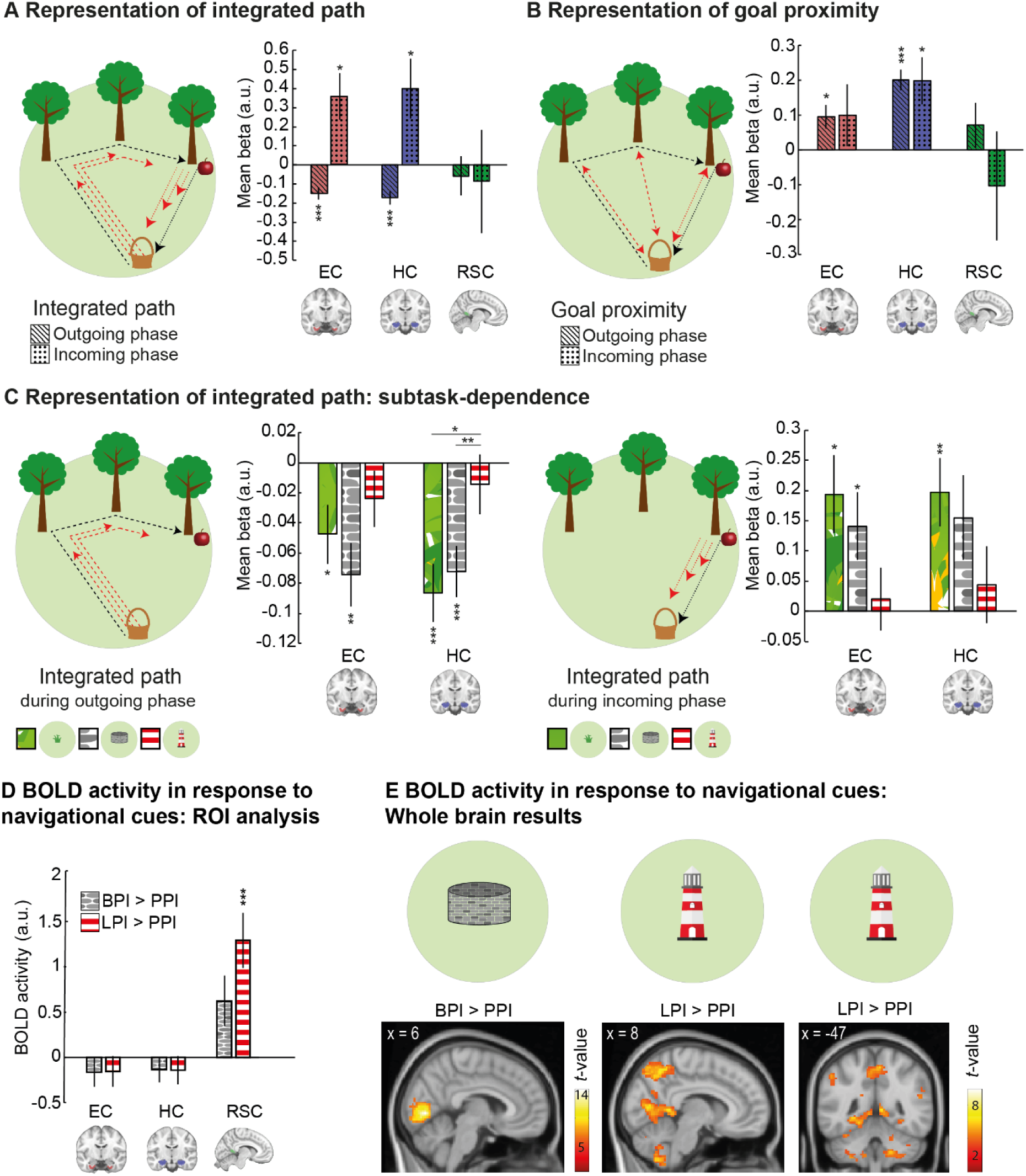
FMRI results: Neural determinants of integrated path, goal proximity, and subtask. (**A**) Representation of integrated path in EC and HC: deactivation during navigation at longer integrated paths during outgoing phase, activation during incoming phase. (**B**) Representation of goal proximity: activation for navigation closer to the goal in HC in both trial phases, and in EC during the outgoing phase. (**C**) Subtask dependence of integrated path representations: HC deactivation during navigation at longer integrated paths during the outgoing phase was modulated by subtask. During the incoming phase, we observed a statistical trend for a similar result in EC and HC. (**D**) Significantly higher activity in RSC during the LPI as compared to the PPI subtask. (**E**) Confirmatory whole brain analyses: Left: activation of visual areas during the BPI as compared to the PPI subtask. Right: higher RSC activity in the LPI than in the PPI subtask. *P*-values are FDR-corrected for the number of contrasts and for the number of ROIs in all ROI analyses (**A, B, D**). For post-hoc comparisons between subtasks in (**C**), *P*-values are FDR-corrected for number of subtasks. \**P* < 0.05; \*\**P* < 0.01; \*\*\**P* < 0.001. Error bars (**A, B, C, D**), s.e.m.; statistical parametric maps in (**E**) are thresholded at *P* < 0.05, FWE corrected for whole brain (left) and small volume corrected for RSC (middle and right), and clusters are considered significant at *P* < 0.05, FWE corrected. FDR, false discovery rate; PI, path integration; PPI, pure path integration; BPI, boundary-supported path integration; LPI, landmark-supported path integration; EC, entorhinal cortex; HC, hippocampus; RSC, retrosplenial cortex; BOLD, blood oxygenation level dependent; a.u., arbitrary units; vm, virtual meters. See also Figures S6, S7, Table S4.

During both the outgoing and the incoming phase, HC activity increased with goal proximity (both *t*_34_ ≥ 2.96, both *P*_FDR_ ≤ 0.028, **Figure 4B**). In EC, we observed this effect only during the outgoing (*t*_34_ = 2.86, *P*_FDR_ = 0.029), but not during the incoming phase (*t*_34_ = 1.12, *P*_FDR_ = 0.813). No relationship between goal proximity and RSC activity was observed.

As integrated path and goal proximity are typically related in the real world, and strongly correlated in our task as well (ρ = -0.38, *Z* = 3.72, *P* < 0.001), we included them in two separate GLMs. We conducted a control analysis to demonstrate that representations of integrated path were not simply side effects of goal proximity representations by including both parametric modulators into a single GLM (Methods). Indeed, results were highly similar (**Supplementary results**).

Together, these results show that BOLD responses in EC and HC represent integrated path and goal proximity, two related variables that are crucial for solving PI tasks. This is in accordance with neurophysiological studies in bats (Sarel *et al.*, 2017) and fMRI studies in humans (Howard *et al.*, 2014; Shine *et al.*, 2019) showing representations of goal direction and distance in EC and HC.

### FMRI correlates of spatial information from boundaries and landmarks

Next, we analyzed which brain regions are recruited by supportive spatial cues. We used a GLM with “task phase” (start phase, outgoing phase, incoming phase, feedback) and “subtask” (PPI, BPI, LPI) as regressors (**Figure S7**) and contrasted activity during the PPI subtask with activity during the BPI or the LPI subtasks. In RSC, activity increased during the LPI as compared to the PPI subtask (*t*_34_ = 4.27, *P*_FDR_ < 0.001; **Figure 4D**), and by trend for the BPI vs. the PPI subtask (*t*_34_ = 2.24, *P*_FDR_ = 0.095). Activity in EC and HC did not differ between the subtasks (all *t*_34_ ≤ 0.97, all *P*_FDR_ ≥ 0.385).

An exploratory whole brain analysis confirmed these results and additionally showed higher activity in V1 and adjacent visual areas during the BPI and LPI subtasks, probably due to the higher amount of visual information (**Figure 4E**; for all clusters, see **Table S4**). Hence, RSC appears to be specifically involved in processing spatial information derived from environmental landmarks, in line with previous results (Mitchell *et al.*, 2018).

### FMRI grid-like representations support PI computations

Previous fMRI studies established a hexadirectional modulation of BOLD activity in the EC as a macroscopic signature of grid cell activity (Doeller, Barry and Burgess, 2010; Kunz *et al.*, 2015, 2019; Bellmund *et al.*, 2016). We thus analyzed GLRs in our paradigm, using a multi-voxel pattern approach (Bellmund *et al.*, 2016) (Methods). We focused on the posterior medial EC (pmEC), the putative human homologue of the rodent medial EC (Hafting *et al.*, 2005; Navarro Schröder *et al.*, 2015; Bellmund *et al.*, 2016). The analysis is based on the rationale that the hexadirectionally symmetric firing pattern of grid cells results in higher pattern similarity when movement directions are 60°, 120°, 180° etc. apart from each other than when they have an offset of 30°, 90°, 150° etc. (**Figure 5A**).

**Figure 5.**
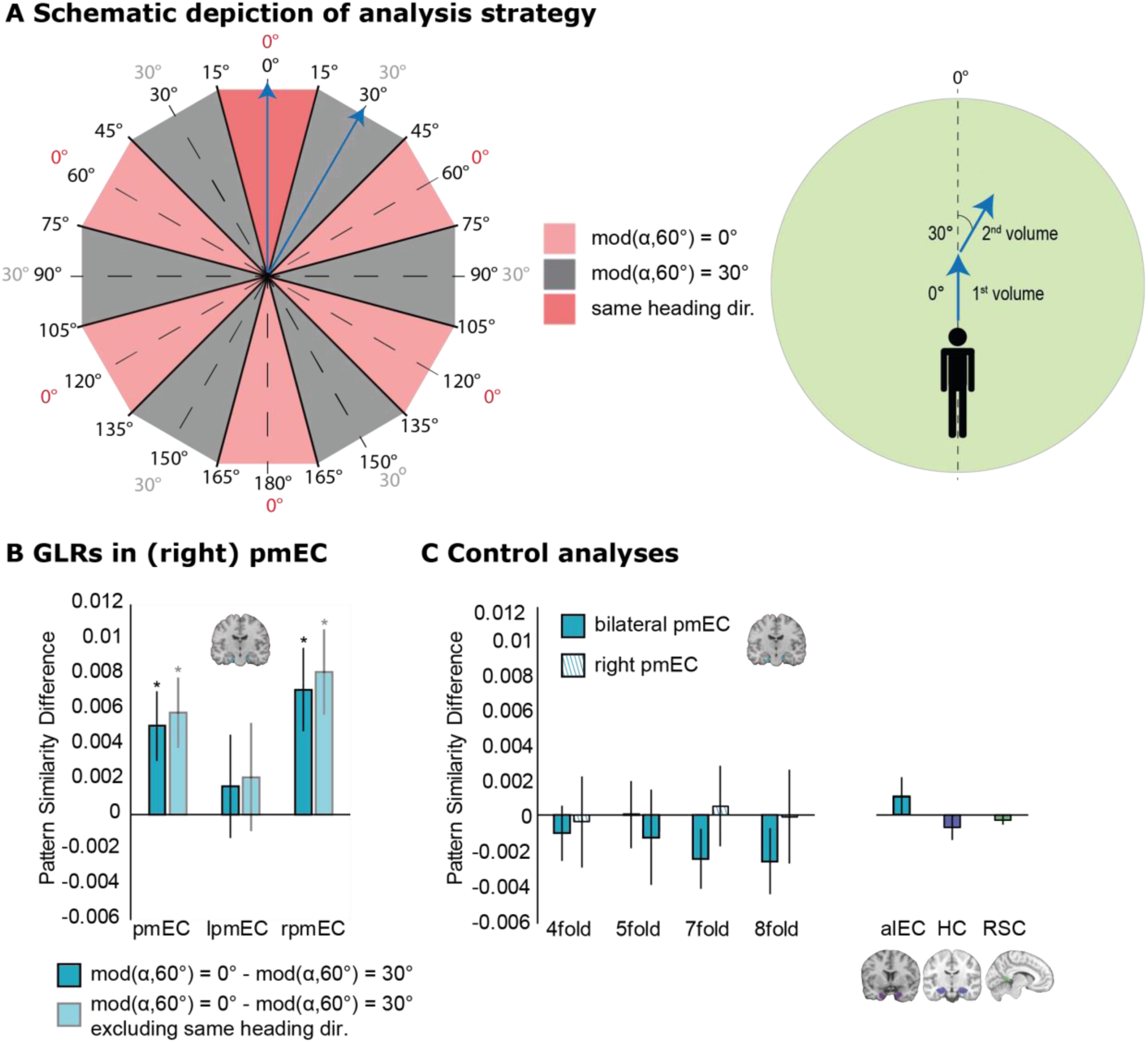
Grid-like representations in pmEC. (**A**) Left: schematic depiction of angular differences in 360° space (inner numbers) and in 60° space (outer numbers). We expected higher pattern similarity for angular differences of mod(α,60°)=0° (rose, where α is the angular difference between two movement directions) as compared to angular differences of mod(α,60°)=30° (gray). We expected the same result when excluding pattern similarities of the same heading direction (dark rose). Right: movement directions of two exemplary fMRI volumes (blue arrows). In the first volume, the subject navigates at an angle of 0° with respect to the reference axis. In the second volume, the subject navigates at an angle of 30°. This results in an angular difference of 30° (in 360° and in 60° space) and thus mod(α,60°)=30° (compare to blue arrows on the left). (**B**) In bilateral and in right pmEC, pattern similarity for angular differences of mod(α,60°)=0° was significantly higher than pattern similarity for angular differences of mod(α,60°)=30°, suggesting a hexadirectional symmetry of pattern similarities (dark blue bars). Same result when removing movements with similar heading directions in 360° space (light blue bars). (**C**) Control analyses. Left: no evidence for 4-fold, 5-fold, 7-fold, or 8-fold rotational symmetry of pattern similarity in bilateral or right pmEC. Right: no evidence for six-fold rotational symmetry of pattern similarity in alEC, HC, and RSC. Error bars (**B, C**), s.e.m. \**P* < 0.05; mod, modulus; dir., direction; GLRs, grid-like representations; pmEC, posterior-medial entorhinal cortex; alEC, anterior-lateral entorhinal cortex; HC, hippocampus; RSC, retrosplenial cortex.

Indeed, we observed a significant hexadirectional symmetry of pattern similarity in pmEC (*t*_34_ = 2.56, *P* = 0.015, **Figure 5B**). A post-hoc test revealed that the effect was confined to right pmEC (*t*_34_ = 2.98, *P* = 0.005), in accordance with previous studies on GLRs (Doeller, Barry and Burgess, 2010; Kunz *et al.*, 2015). GLRs were not driven by a head direction signal: Eliminating movement directions with angular differences of -15° to 15° still resulted in a hexadirectional signal (bilateral pmEC: *t*_34_ = 2.90, *P* = 0.006; right pmEC: *t*_34_ = 3.36, *P* = 0.002, **Figure 5B**). We performed a series of control analyses to ensure validity and specificity of the GLRs (**Supplementary Results, Figure 5C**).

### Neural representations of spatial features predict PI performance

Finally, we aimed at establishing a mechanistic model that could explain PI performance (**Figure 6**). We built an exploratory mixed linear model with “subtask” and “incoming distance” as within-subject predictors, “subject” as random factor, and the following fMRI-based between-subjects predictors: (i) EC representations of integrated path during the incoming phase, (ii) HC representations of goal proximity during the incoming phase (iii) GLRs in pmEC, and (iv) RSC landmark representations (Model 6; **Table 1**). The other potential fMRI-based predictors were not included into the model as they either caused multicollinearity or because they did not significantly improve model fit (for a detailed explanation of the model build-up, see Methods). We allowed for interactions of the fMRI-based predictors with subtask or incoming distance, but not with the other fMRI-based predictors. Post-hoc comparisons were Tukey-corrected for multiple comparisons (number of subtasks or number of compared quintiles).

**Figure 6.**
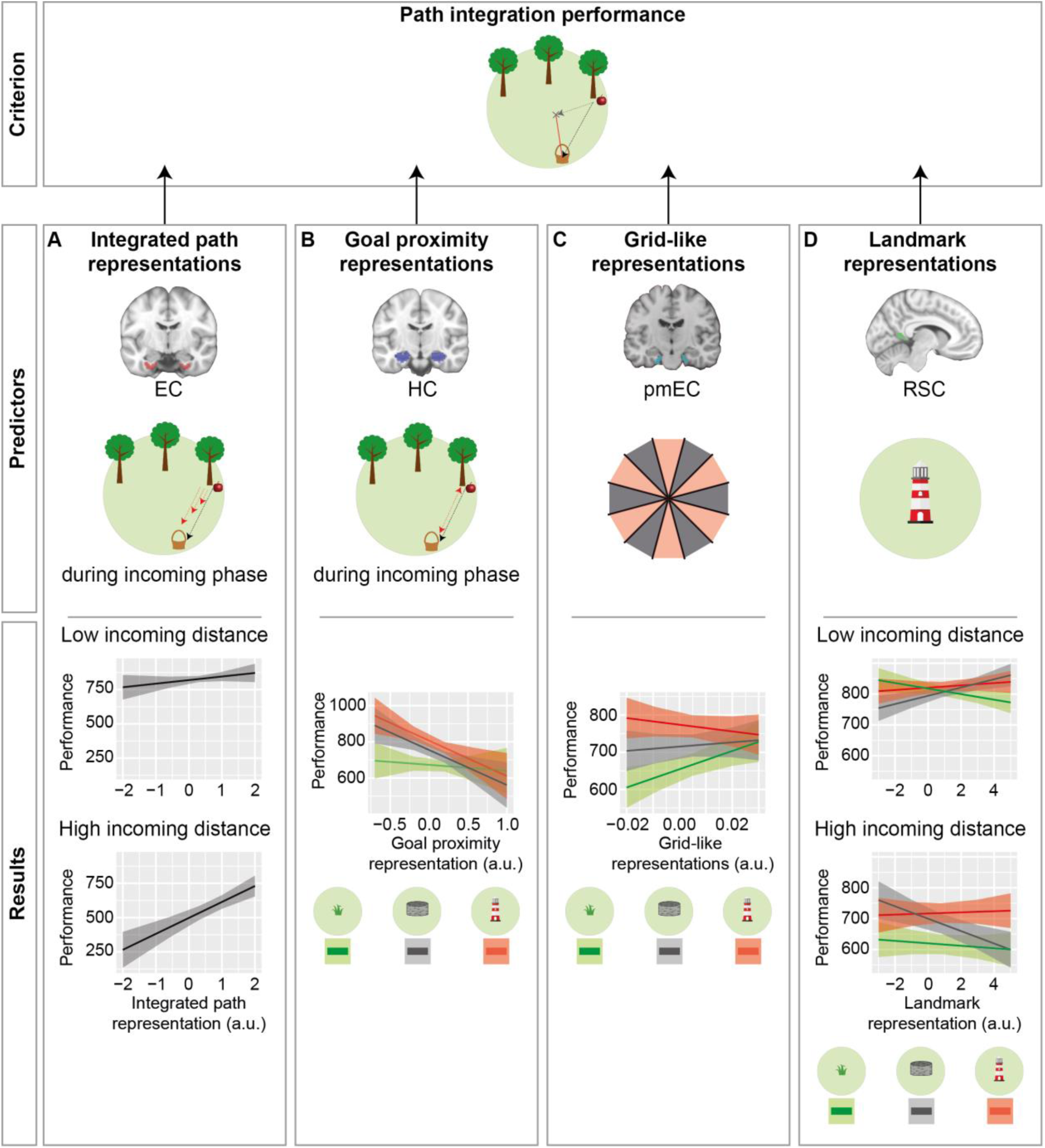
Mechanistic model to predict PI performance. We aimed at predicting PI performance as a function of fMRI-based representations of spatial features in combination with subtask and incoming distance (model 6). (**A**) Integrated path representations in EC interacted with incoming distance in predicting PI performance. At higher incoming distances, stronger integrated path representations in EC were associated with better performance. (**B**) Goal proximity representations in HC interacted with subtask in predicting PI performance. In none of the subtasks was the prediction significant by itself. (**C**) GLRs in pmEC interacted with subtask in predicting PI performance. Only in PPI, higher GLRs in pmEC were associated with better performance. (**D**) Landmark representations in RSC interacted with subtask and incoming distance in predicting PI performance. In none of the individual subtask by incoming distance combinations was the prediction significant by itself. As the model contained two continuous predictors, one of them (incoming distance) was discretized into quintiles for post-hoc tests and for graphical depiction; only quintiles 1 and 5 are depicted (**A, D**). Y-axes show parameter estimates for performance; shaded areas, s.e.m; PI, path integration; PPI, pure path integration; BPI, boundary-supported path integration; LPI, landmark-supported path integration; EC, entorhinal cortex; HC, hippocampus; pmEC, posterior-medial EC; RSC, retrosplenial cortex; a.u., arbitrary units; vm, virtual meters.

First, we found that EC representations of integrated path during the incoming phase predicted performance (*F* = 16.19, *P* < 0.001). This effect was more pronounced at high incoming distances, as shown by a significant interaction with incoming distance (*F* = 5.43, *P* = 0.020): At higher incoming distances (quintiles 2–5), EC representations of integrated path showed a significant association with performance (all *z* ≥ 3.32, all *P*_Tukey_ < 0.005). This was not the case at low incoming distances (quintile 1, *z* = 1.97, *P*_Tukey_ = 0.221).

Second, while we did not encounter a significant main effect of HC representations of goal proximity during the incoming phase (*F* = 1.39, *P* = 0.247), we found a significant interaction with subtask (*F* = 3.43, *P* = 0.032). However, post-hoc tests revealed no significant association between this fMRI predictor and performance in any subtask (all *z* ≤ 2.08, all *P*_Tukey_ ≥ 0.109).

Third, the magnitude of GLRs had no main effect on performance (*F* = 0.73, *P* = 0.398), but showed a significant interaction with subtask (*F* = 7.98, *P* < 0.001). Post-hoc comparisons revealed that GLRs predicted performance to a greater extent in PPI than in LPI (*z* = 3.99, *P*_Tukey_ < 0.001). No differences were encountered between the other subtask combinations (both *z* ≤ 2.14, both *P*_Tukey_ ≥ 0.082). When analyzed separately, we found that GLRs significantly predicted performance in the PPI subtask (*z* = 2.52, *P*_Tukey_ = 0.035), but not in the other subtasks (both *z* ≤ 0.96, both *P*_Tukey_ ≥ 0.711). These results indicate that the grid cell system in pmEC supports PI specifically in the absence of supportive spatial cues.

Finally, the magnitude of RSC representations of landmarks showed no main effect on performance (*F* = 0.07, *P* = 0.793), but we encountered a significant two-way interaction with subtask (*F* = 6.93, *P* = 0.001) and a three-way interaction with subtask and incoming distance (*F* = 4.36, *P* = 0.013). However, RSC representations of landmarks predicted performance in none of the individual follow-up tests (all *z* ≤ 1.47, all *P*_Tukey_ ≥ 0.543).

## Discussion

Our behavioral results show that *APOE* ε4-carriers perform as well as controls as long as they can use supportive spatial information from a boundary or a landmark. Their deficit in PI is unmasked, however, when potentials for error accumulation increase (i.e., with higher PI distance) and/or when no compensatory strategies can be employed. In these conditions, PI performance relates to EC gray matter volume and to the strength of entorhinal GLRs. Supportive spatial cues improve performance, presumably via compensatory recruitment of other regions including the RSC.

We propose that changes in grid cell functioning underlie the impaired performance of risk carriers during pure PI because various studies identified grid cells as the main neural substrate for PI (Hafting *et al.*, 2005; O’Keefe and Burgess, 2005; Burak and Fiete, 2009; Kubie and Fenton, 2012; Banino *et al.*, 2018; Gil *et al.*, 2018; Stangl *et al.*, 2018). Indeed, in our fMRI study we observed a positive relationship between the strength of GLRs and PI performance exclusively in the absence of supportive spatial cues that could reduce error accumulation during PI computations (Gallistel, 1990; Burak and Fiete, 2009; Hardcastle, Ganguli and Giocomo, 2015). This indicates that the experimental condition in which risk carriers showed impaired PI performance indeed relied most strongly on the grid cell system. Risk carriers may thus experience higher error accumulation during pure PI due to a compromised EC grid cell system.

Furthermore, we found representations of integrated path and goal proximity in EC and HC during the outgoing as well as during the incoming phase. This result is in line with models suggesting that EC grid cells provide the computational basis for representing the integrated path (i.e., the distance and direction traveled) (Burak and Fiete, 2009). A complementary model suggests that grid cells enable the computation of a direct vector to the goal (O’Keefe and Burgess, 2005; Kubie and Fenton, 2012; Howard *et al.*, 2014; Epstein *et al.*, 2017). Accordingly, we demonstrated that risk carriers’ performance is particularly affected by high incoming distances, and that this is associated with a higher dependency on EC volume in *APOE* ε4-carriers.

An environmental boundary or a landmark allowed risk carriers to perform as well as controls. This is consistent with a recent study showing similar PI performance in risk and control participants in rich virtual environments (Coughlan *et al.*, 2019). Using fMRI, we demonstrated increased RSC activity in response to an environmental landmark, consistent with previous studies (Mitchell *et al.*, 2018). The RSC provides a neural basis for viewpoint-dependent representations of local place and direction (Marchette *et al.*, 2014), which may be at the core of compensatory strategies employed by risk carriers. We propose that risk carriers recruited the RSC in the landmark condition to a greater extent than controls in order to counteract error accumulation during PI and to compensate for a reduced reliability of GLRs, thus anchoring the cognitive map (Epstein, 2008; Epstein *et al.*, 2017). In support of this idea, risk carriers showed a stronger relationship between goal-to-landmark distance and performance, and navigated in closer proximity to the landmark during the incoming phase. However, future fMRI studies with genotyped participants are required to corroborate this idea.

In the BPI subtask, the relationship between performance and goal-to-boundary distance was not modulated by genotype, and risk carriers did not navigate closer to the boundary as compared to control participants. This null finding differs from previous findings showing preferred navigation of risk carriers along environmental boundaries (Kunz *et al.*, 2015; Coughlan *et al.*, 2019). This discrepancy may be explained by the specific layout of our BPI subtask: the boundary was at considerable distance to potential goal locations, and navigating towards the boundary led to a linear decrease in navigation speed. The exact effects of environmental boundaries on PI performance may be scrutinized in future studies by comparing different types of boundaries (squared, circular) and environmental layouts (distal and/or proximal landmarks).

Compensatory strategies of EC-dependent behavioral deficits in *APOE* ε4-carriers may be relevant for the progression of AD pathology: It has been suggested that neural hyperactivity associated with these strategies leads to a progressive deterioration in structural integrity of relevant brain areas (Dickerson *et al.*, 2005; Ewers *et al.*, 2011), perhaps by increasing amyloid-β (Jagust and Mormino, 2011) and/or tau deposition (Huijbers *et al.*, 2019). Thus, potential compensatory strategies in risk carriers associated with higher activity in specific brain areas may contribute to AD progression (Kunz *et al.*, 2015). Consequently, at later stages of AD, patients should no longer be able to rely on compensatory mechanisms (Prieto del Val, Cantero and Atienza, 2015), leading to worse performance irrespective of whether supportive spatial cues are provided. Indeed, in a recent study, mild cognitive impairment patients with positive AD biomarkers showed reduced PI performance irrespective of the availability of supportive spatial cues (Howett *et al.*, 2019). At this stage, neural hyperactivity may even directly exert detrimental effects on behavior (Bakker *et al.*, 2012). Preventing hyperactivity might thus be a promising therapeutic strategy, possibly improving interneuron dysfunction and counteracting network abnormalities (Palop and Mucke, 2016).

## Conclusion

EC-dependent functioning may prove particularly useful for predicting AD because of its early affection by AD-related neuropathological changes (Braak *et al.*, 2011). Distinct navigational strategies may deteriorate at different preclinical and clinical stages of AD, and pure PI may capture the behavioral manifestation of earliest neuropathological changes related to preclinical AD development.

## Data Availability

Data available on request from corresponding authors.

## Acknowledgements

We are grateful to all participants who volunteered to participate in this study. We thank A. Bäumer, V. Ruzas, R. Schütte, and S. Wetter for help with behavioral data acquisition at IfADo; S.M. Cammisuli for technical support and S. Squitieri for recruitment planning and administrative procedures at the Italian site; E. Genc for advice with sMRI data analysis; and T. Navarro Schröder for providing the ROI mask of the pmEC. The acquisition of participants and the *APOE* genotyping at IfADo were carried out as part of the Dortmund Vital Study. L.K. was supported by the BMBF (01GQ1705A), NIH grant 563386, NSF grant BCS-1724243, and the BrainLinks-BrainTools Cluster of Excellence funded by the German Research Foundation (DFG, EXC 1086). A.P. and F.B were partially supported by Emma and Ernesto Rulfo Foundation for Medical Genetics. J.L.C. and M.A were supported by research grants from the Spanish Ministry of Economy and Competitiveness (SAF2017-85310-R to JLC, PSI2017-85311-P to MA); the Regional Ministry of Innovation, Science and Enterprise, Junta de Andalucia (P12-CTS-2327 to JLC); the International Center on Aging CENIE-POCTEP (0348_CIE_6_E to MA); and CIBERNED (JLC). N.A. received funding by the Deutsche Forschungsgemeinschaft (DFG, German Research Foundation) – Projektnummer 316803389 – SFB 1280 as well as via Projektnummer 122679504 – SFB 874.

## Author contributions

L.K. and N.A. developed key hypotheses. A.B., L.K., and N.A. designed the paradigm. A.B., L.K., S.G., P.D.G., E.W., D.M.C., J.L.C., and N.A. designed the study. A.B., C.A.G., S.G., E.W., P.D.G., D.M.C., F.B., C.P., A.P., Y.B., and J.L.C. collected data. A.B., L.K., C.A.G., and N.A. analyzed data with support from M.L. and L.D. A.B., L.K., and N.A. wrote the manuscript, with substantial support from all authors.

## Methods

### Participants

We recruited healthy participants at five European sites including Germany (2 sites), Spain, Belgium, and Italy (**Table S1**) to (i) examine the influence of *APOE* genotype on PI performance (“*APOE* sample”); (ii) elucidate the fMRI signatures of PI in our task (“fMRI sample”); and (iii) investigate associations between brain structure and PI performance (“sMRI sample”). The study was performed in accordance with the declaration of Helsinki and was approved by the respective institutional review boards at all sites. All participants gave their written informed consent.

### APOE *sample*

Four groups of participants (total, *N* = 318) were recruited at four different sites: in Germany (“*APOE* sample 1”; *n* = 112; IfADo – Leibniz Research Centre for Working Environment and Human Factors at the Technical University Dortmund, Dortmund, Germany), Spain (“*APOE* sample 2”; *n* = 114; Pablo de Olavide University, Seville, Spain), Italy (“*APOE* sample 3”; *n* = 68; University of Parma, Parma, Italy) and Belgium (“*APOE* sample 4”; *n* = 24; Cliniques Universitaires Saint-Luc, Brussels, Belgium). 51 participants were excluded because of genotypes other than ε3/ε4 or ε3/ε3 (*n* = 46), prior familiarity with the task (*n* = 1), or technical reasons (*n* = 4), resulting in a final sample of *n* = 267 participants (for demographics, see **Table S1, Figures S2, S3**).

### SMRI sample

The sMRI sample consisted of a subset of participants from *APOE* sample 2 for which sMRI scans were available (*n* = 99; for demographics, see **Table S1, Figure S3)**.

### FMRI *sample*

To elucidate the fMRI signatures of PI in our task, we recruited young healthy participants (*n* = 35; for demographics, see **Table S1**) at Ruhr University Bochum, Bochum, Germany.

### APOE *genotyping*

Four groups of participants (*APOE* samples 1-4) were analyzed for the *APOE* polymorphisms rs429358, a [C/T] substitution on chromosome 19q13.32 of the sequence GCTGGGCGCGGACATGGAGGACGTG[C/T]GCGGCCGCCTGGTGCAGTACCGCGG and rs7412, a [C/T] substitution of the sequence CCGCGATGCCGATGACCTGCAGAAG[C/T]GCCTGGCAGTGTACCAGGCCGGGGC. Based on the two single nucleotide polymorphisms participants were assigned to one of the three alleles ε2, ε3 or ε4 as described before (Zhong *et al.*, 2016).

For *APOE* sample 1, venous blood was taken and DNA was isolated using a QIAamp DNA blood maxi kit (Qiagen, Hilden, Germany) according to the manufacturer’s protocol (Arand *et al.*, 1996). DNA concentrations were determined using a NanoDrop ND-1000 UV/Vis-spectrophotometer (PEQLAB Biotechnologie GMBH, Erlangen, Germany). Genotyping was performed on an ABI7500 Sequence Detection System with the use of TaqMan assays (assay ID: C_3084793_20 for rs429358; assay ID: C_904973_10 for rs7412; Applied Biosystems, Darmstadt, Germany). Analysis of data was performed according to the manufacturer’s instructions (Applied Biosystems 7300/7500/7500, fast real-time PCR System Allelic Discrimination Getting Started Guide).

For *APOE* sample 2, genomic DNA was isolated from blood using a standard salting-out protocol (Miller, Dykes and Polesky, 1988). DNA concentration and purity was determined by UV spectrophotometric measurements (Quawell, Q3000 UV). Genotyping was performed by real time PCR (Step-One Plus, Applied Biosystem) using pre-designed TaqMan SNP genotyping assays (Assay ID: C_3084793_20 for rs429358; Assay ID: C_904973_10 for rs7412; Applied Biosystems, Darmstadt, Germany).

For *APOE* sample 3, genomic DNA was extracted from buccal brushs using the Gentra Puregene Buccal Cell Kit (Qiagen, Valencia, CA, USA). The *APOE* SNPs were genotyped using an ABI PRISM 7700 Sequence Detector (assay ID: C_3084793_20 for rs429358; assay ID: C_904973_10 for rs7412; Thermo Fisher Scientific, MA, USA) with a TaqMan 5′-allele discrimination Assay-By-Design method (Thermo Fisher Scientific, MA, USA). PCR was performed according to the manufacturer’s instructions.

For *APOE* sample 4, DNA was extracted from blood and analysis of *APOE* polymorphisms was performed by means of restriction enzyme isoform genotyping (Hixson and Vernier, 1990). We used pre-designed TaqMan SNP genotyping assays (assay ID: C_3084793_20 for rs429358; assay ID: C_904973_10 for rs7412; Applied Biosystems, Darmstadt, Germany). Participants were not genetically preselected. For analyses, we focused on two genetic subgroups: *APOE* ε3/ε3 carriers [“control group”; common genetic risk for AD (Corder *et al.*, 1993); *n* = 202] and *APOE* ε3/ε4 carriers [“risk group”; increased genetic risk for AD (Corder *et al.*, 1993); *n* = 65]. Risk group and control group did not differ in terms of demographic characteristics (**Table S2** for the *APOE* sample and **Table S3** for the sMRI sample). Participants from other genetic subgroups were excluded as in previous studies (Kunz *et al.*, 2015, 2017) (ε2/ε2, *n* = 2; ε2/ε3, *n* = 33; ε2/ε4, *n* = 5; ε4/ε4, *n* = 5; for genotype distributions in the subsamples, see **Table S5**). Participants as well as experimenters were blinded towards genotypes. Sample size was based on previous studies, suggesting >50 participants in the smaller genetic subgroup (Shaw *et al.*, 2007; Ruiz *et al.*, 2010; Nichols *et al.*, 2012; Matura *et al.*, 2014; Zhang *et al.*, 2015; see also Weissberger *et al.*, 2018). All participants reported normal or corrected-to-normal vision and no history of neurological or psychiatric diseases.

### Experimental task

Participants performed a PI task (the “Apple Game”) in a virtual environment implemented via Unreal Engine (Epic Games, version 4.11). The environment consisted of an endless grassy plane with a blue sky rendered at infinity. Trials were divided into three phases. During the “start phase”, participants navigated to a “goal location” which was marked by a basket (**Figures 1B**,**C**). Participants were instructed to remember this location. When they had reached the goal location, the basket disappeared and a tree appeared in a different location (“outgoing phase”). Participants walked towards the tree, checked if it had an apple, and walked to the tree to make it disappear. If the tree did not have an apple, another tree appeared. Trials contained between 1 and 5 trees to systematically vary PI difficulty, corresponding to a variation of “outgoing distance” (**Figure 1C**, see also “Behavioral analyses”; for similar tasks, see Loomis *et al.*, 1993; Tcheang, Bulthoff and Burgess, 2011; Mokrisova *et al.*, 2016; Stangl *et al.*, 2018; Coughlan *et al.*, 2019; Howett *et al.*, 2019). The final tree was marked by a red apple (“retrieval location”), indicating that the participants had to navigate back to the goal location (to place the apple into the basket; “incoming phase”). After pressing a button to indicate that this was the presumed location of the basket (“response location”), participants received feedback via zero to three stars, depending on the Euclidean distance between the response location and the correct goal location (“drop error”; **Figure 1D**; <1600 vm for 3 stars, <3200 vm for 2 stars, <6400 vm for 1 star; **Figure S1**). All phases were self-paced and only the “incoming phase” had a time limit of 60 seconds before the next trial started (this time limit was never reached by any participant). Locations of baskets and trees were equally distributed across an (invisible) grid of 8×8 squares (bin edge length, 800 vm) such that each participant visited all squares once in each environmental condition (**Figure S1**).

PI performance was tested in three environmental conditions that differed with regard to the presence or absence of supportive spatial cues. In the “pure PI” (PPI) condition, the virtual environment did not contain any landmarks or boundaries, and participants purely relied on visual flow to perform PI (**Figure 1A**). In the “boundary-supported PI” (BPI) condition, a circular stonewall with a height of 2050 vm surrounded the environment at a radius of 6788 vm (**Figure 1A**, middle; for location and radius see also **Figure S1**). In the “landmark-supported PI” (LPI) condition, a lighthouse with a height of 1300 vm was present at × = 1600 vm and y = 800 vm, serving as an intramaze landmark (**Figure 1A**, for the location see also **Figure S1**). No distal cues outside the boundary were present, which is different from previous studies examining the influence of *APOE* on navigational behavior (Kunz *et al.*, 2015; Coughlan *et al.*, 2019). We chose this design in order to specifically assess the effects of the boundary and the landmark.

Participants navigated the virtual environments using a joystick (behavioral experiment: Trust GXT 555 Predator; fMRI experiment: MR-compatible joystick from NAtA-Technologies, Coquitlam, Canada), allowing them to move forward, turn left, or turn right. Moving backwards was not possible so that movement direction was equivalent with heading direction. In each subtask, participants’ speed was attenuated when their distance from the center of the arena was larger than 5657 vm and linearly decreased to zero at 6788 vm ensuring a constant movement radius in subtasks with and without a visible boundary (**Figure S1**). In this “speed reduction zone”, participants could navigate at full speed when heading towards the center of the arena. The position of the participant was logged every 200 ms, which allowed us to extract movement periods, movement speed, and movement direction.

### Behavioral Experiment

The paradigm was subdivided into subtasks of 16 trials each. Subtasks varied with respect to the layout of the virtual environment. Participants started the first trial of each subtask in the center of the virtual environment (x = 0 vm, y = 0 vm). The outgoing phase of each subtask contained either 1 tree (3 trials), 2 trees (3 trials), 3 trees (4 trials), 4 trees (3 trials), or 5 trees (3 trials) in randomized order (including the tree marked by an apple). This trial procedure allowed for a perfect balancing of locations during the experiment, so that each participant visited all 64 squares once in each subtask (**Figure S1**). Before the beginning of the task, all participants completed eight trials in the PPI condition in order to practice the paradigm. As we encountered systematic within-subject effects of fixed trial and location sequences in a subgroup of participants (for the exact number of participants, refer to **Table S2**), we fully randomized trials and locations for later participants. The randomization neither showed a main effect on performance (*F* = 0.74, *P* = 0.477), nor did any of the results change when we added randomization version as a covariate to the model (“Subtask”: *F* = 799.09, *P* < 0.001; “Incoming distance”: *F* = 4139.47, *P* < 0.001; “*APOE* × Subtask”: *F* = 10.89, *P* < 0.001; performance difference in PPI between risk carriers and controls: *z* = 2.08, *P*_Tukey_ = 0.037). Participants in Parma, Seville, and 20 participants in Brussels completed a long version of the paradigm with six subtasks (2 x PPI, 2 x BPI, and 2 x LPI; **Figure 1E**), with the order of the subtasks being pseudorandomized such that the same subtask would not follow each other and the three different subtasks would be equally distributed across the experimental halves. This resulted in a total of 96 experimental trials. Participants could take breaks between the subtasks. The experiment lasted 107.44 ± 33.15 min [mean ± standard deviation (s.d.)] in total. Participants in Dortmund and four participants in Brussels completed a shorter version of the paradigm with three subtasks (1 x PPI, 1 x BPI, and 1 x LPI; **Figure 1E**) with the order of the subtasks being randomized resulting in 48 experimental trials. Including breaks between the subtasks and practice trials, the short version of the experiment lasted 52.83 ± 12.08 min (mean ± s.d.) in total.

### FMRI experiment

The fMRI experiment consisted of a practice run, which was conducted during the structural scan (6 min), and two functional runs. The practice run comprised a maximum of nine trials (three trials in each condition) and ended when the structural scan was over. Each functional run had a duration of 22.32 ± 3.18 min (mean ± s.d.), which corresponds to 536 ± 76 (mean ± s.d.) fMRI volumes. The duration of the functional runs varied between participants, since the entire task was self-paced. In-between runs, participants could take short breaks. Each run consisted of six blocks containing four trials of each of the three environmental conditions (**Figure 1F**), resulting in 16 trials per condition and thus 48 trials in total across the experiment. The order of the blocks was pseudorandomized such that the same subtask would not follow each other and the three different subtasks would be equally distributed across the two experimental runs. Before every new trial, participants viewed a fixation crosshair with a variable duration of 5 to 7.5 s (randomly distributed).

### Data acquisition

Data was acquired according to a standardized protocol (with regard to experimental setup, instructions, behavior of the investigator, and breaks), ensuring comparability across the five recording sites.

### *Behavioral data acquisition (*APOE *sample)*

The paradigm was presented on laptops with a screen diagonal of 45 cm, a resolution of 1920 x 1080 pixels and a frame rate of 60 frames per second. We attached the joystick to the table by means of a custom-made frame (identical for all sites), which also served as an armrest and placed the laptop at a distance of 50 cm in front of the participant.

### MRI data acquisition (sMRI sample)

Structural brain images were acquired at the Neuroimaging Service of the Pablo de Olavide University (Seville, Spain) using a 3T Philips Ingenia CX MRI scanner equipped with a 32-channel receiver head coil (Philips, Best, Netherlands). Head motion was minimized by placing foam padding around the subject’s head. One high-resolution three-dimensional (3D) T1-weighted Magnetization-Prepared Rapid Gradient Echo (MP-RAGE) sequence was acquired in the sagittal plane. Acquisition parameters were empirically optimized to enhance the gray/white matter contrast: TR/TE = 2600/4.7 ms, flip angle (FA) = 9°, voxel resolution = 0.65 mm isotropic, acquisition matrix = 384 x 384 mm, resulting in 282 contiguous slices without gap between adjacent slices, acceleration factor (SENSE) = 1.7, and field of view (FOV) = 250 x 250 x 183 mm^3^.

### fMRI data acquisition (fMRI sample)

The fMRI recordings were conducted at the Bergmannsheil hospital in Bochum using a 3T Philips Achieva scanner (Best, The Netherlands) with a 32-channel head coil. High-resolution whole-brain structural brain scans of participants were acquired using a T1-weighted sequence at 1 mm isotropic resolution, a field of view (FOV) of 240 x 240 mm^2^, and 220 transversally oriented slices during a total acquisition time (TA) of 6 min 2 s. Blood oxygenation level dependent (BOLD) contrast images were measured with a T2* weighted gradient echo EPI sequence with the following parameters: 2.5 mm isotropic resolution, repetition time (TR) = 2500 ms, echo time (TE) = 30 ms, flip angle (FA) = 90°, FOV = 96 mm x 96 mm, 46 transversal slices in interleaved order without slice gap, TA = 22.32 ± 3.18 min (mean ± s.d.), corresponding to 536 ± 76 volumes (mean ± s.d.). We discarded the first five images of each session to allow for signal steady-state transition. Participants viewed the virtual environment via MR-compatible LCD-goggles (Visuastim Digital, Resonance Technology Inc., Northridge, CA, USA) with a resolution of 800 x 600 pixels and they navigated the virtual environment by means of an MR-compatible joystick.

### Data analysis

We extracted behavioral data from logfiles using Matlab (2018a, TheMathWorks Inc., Massachusetts) including the Parallel Computing Toolbox (v6.12) and the CircStat Toolbox (Berens, 2009). Regions of interest (ROIs) were created using FreeSurfer (v6.0.0). We used SPM12 (http://www.fil.ion.ucl.ac.uk/spm) for all fMRI analyses. Statistics were done in R (R Core Team, 2018; 3.5.0), using the lme4 (Bates *et al.*, 2014; v1.1-17) and the emmeans (Lenth, 2019; v1.2.2) packages.

### Behavioral analyses

We determined three different performance measures from the behavioral logfiles (**Figure 1D**): “drop error”, “distance error”, and “rotation error”. The drop error was defined as the Euclidean distance between the goal location and the response location. We computed the distance error as the absolute difference between two distance measures: distance error = D_correct_ - D_response_, where D_correct_ is the distance between the retrieval and the goal location (i.e., the distance of the correct incoming path), and D_response_ is the distance between the retrieval and the response location (i.e., the distance of the incoming path chosen by the participant). Rotation error was computed correspondingly as the absolute difference between two angular measures: rotation error = R_correct_ - R_response_, where R_correct_ is the heading direction of the correct incoming path, and R_response_ is the heading direction of the response path. Only for visualization and enhanced readability, we converted the error measures to performance measures of participant *i* (performancei) using a linear transformation (Eq. 1):

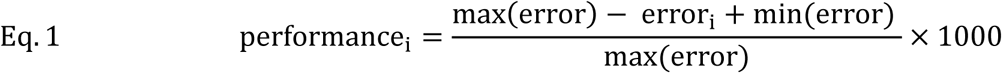

where max(error) and min(error) correspond to the maximum and minimum error across all participants, respectively, and errori corresponds to the error in participant *i*. The formula simply reverses the errors and maps them into the range between 0 and 1000. Note that the linear transformation does not affect the statistical results. Thus, “performance” refers to the drop error, “performance based on distance error” refers to the distance error, and “performance based on rotation error” refers to the rotation error.

In order to analyze performance as a function of path distance, we calculated “incoming” and “outgoing distance” (**Figure 1C**): Outgoing distance was determined as the cumulated path distance during the “outgoing phase”, i.e., we summed the Euclidean distances between “goal”, “tree without apple”, and “retrieval” locations. Incoming distance was defined as the Euclidean distance between the retrieval location and the goal location. Outgoing and incoming distance reflect different sub-components of PI (keeping track of the traveled path with regard to the goal location and computing a straight line in relation to the goal location, respectively), but are putatively both subject to cumulative error accumulation (Gallistel, 1990; Etienne and Jeffery, 2004; Burak and Fiete, 2009; Kubie and Fenton, 2009).

The precision of PI processes can be improved if sensory information is available to recalibrate the ideothetic coding process (Gallistel, 1990; Etienne and Jeffery, 2004; Hardcastle, Ganguli and Giocomo, 2015). We thus investigated the influence of spatial cue distance, i.e., the distance between the goal location and the boundary (“goal-to-boundary” distance) or the landmark (“goal-to-landmark” distance) on performance.

Finally, we examined navigational strategy, irrespective of performance. To this end, we computed the mean Euclidean distance from the boundary or from the landmark across all time points of the incoming phase (this was not computed during the outgoing phase, because navigation during this period was determined by the consecutive trees).

### Statistical analysis

To assess potential demographic differences between risk carriers and controls, we performed two-tailed *t*-tests, Wilcoxon tests, and chi-squared tests. To test for significant relationships between fMRI representations, we performed bivariate Pearson or Spearman correlations. In order to disentangle the various factors driving PI performance in the different subtasks of our paradigm, we implemented a mixed linear model using the lme4 package in R (Bates *et al.*, 2014; R Core Team, 2018). We were both interested in fixed effects of predictors varying within subjects and in effects of predictors varying between subjects.

As within-subject predictors, we included subtask (i.e., PPI, LPI, and BPI) and two different types of distance predictors: a path distance predictor that could be quantified in all three subtasks (i.e., outgoing distance or incoming distance), and a spatial cue distance predictor that was only defined in the corresponding subtask (goal-to-boundary distance in the BPI subtask and goal-to-landmark distance in the LPI subtask). In addition, we considered between-subject effects: genotype (i.e., *APOE* ε4-carriers and *APOE* ε4-noncarriers) that was defined for all subjects in the *APOE* sample; volume in EC, HC, or RSC, that was only defined for subjects with available structural MRI data (sMRI sample); and representations of integrated path, goal proximity, boundary, landmark, and grid-like representations (GLRs), that were only defined for subjects with available fMRI data (fMRI sample).

Continuous variables were centered on the grand mean (between-subject predictors) or on the subject mean (within-subject predictors). We included age and sex as covariates to control for main effects and interactions on subtask, genotype, and gray matter volume. Site and subject were included as random factors. We tested separate models for each outcome variable (performance, performance based on distance error, and performance based on rotation error). Since it is most appropriate for model building and preferable in terms of power (Langsrud, 2003), we always used Type II Sum of Squares (SS) to test fixed effects. For post-hoc pairwise comparisons, we used Tukey-adjusted Fisher’s tests as implemented in the emmeans package (Lenth, 2019). All statistical tests were two-tailed at an alpha level of α < 0.05. We did not report regression coefficients and degrees of freedom, as both are not intuitively interpretable in mixed linear models of high complexity.

We reported all significant main effects and interactions as well as non-significant effects which were of interest for our hypotheses. Thus, effects that are not reported in the results section were not significant. We did not report significant interactions with covariates (but see **Figure 4**).

All mixed linear models, except model 6, were built in a hypothesis-driven way (see **Table 1** for an overview of all models): Models 1a-1b were built to test for effects of subtask, genotype, and the two path distance regressors on performance. We used separate models for the two path distance predictors (“a”, outgoing distance; “b”, incoming distance), because they were positively correlated and would have caused multicollinearity. However, in order to scrutinize whether the two predictors indeed showed significantly different interactions with genotype, we built model 1c. This model contained one regressor that accounted for the effects of the path distance predictors irrespective of the type of path distance, and a binary regressor that accounted for the type of path distance (0 = outgoing distance, 1 = incoming distance).

Models 2a-2b served to analyze the effects of spatial cue distance, i.e., the subtask-specific measures of the distance between the goal and either the landmark (LPI subtask) or the boundary (BPI subtask). In Models 2a-2b, we analyzed the effects of genotype and goal-to-boundary or goal-to-landmark distance on performance in the respective subtasks.

Models 2c-2d were the only models in which we did not test for factors influencing PI performance, but tested whether *APOE* genotype predicted whether subjects navigated closer to the boundary (BPI subtask) or the landmark (LPI subtask), similar to previous studies (Kunz *et al.*, 2015; Coughlan *et al.*, 2019). Thus, we did not use performance as criterion (i.e. dependent variable) but distance-to-boundary (BPI subtask) or distance-to-landmark (LPI subtask), respectively. These distance measures were obtained by averaging the Euclidean distances across all time points of the incoming phase of each trial of the respective subtasks. Models 3-5 tested for effects of EC, HC, and RSC volumes on PI performance, respectively. These between-subject variables were included in addition to genotype, subtask, and one of the path distance measures.

Finally, model 6 served to predict performance by means of incoming distance, subtask and different fMRI representations. As we did not have specific hypotheses about the predictors, this model was built in an exploratory way (Bates *et al.*, 2015) with the restrictions that overfitting and multicollinearity had to be avoided. The following fMRI representations were considered as model predictors, because of significant effects in prior analyses (**Figures 4, 5**):

1. GLRs in bilateral pmEC
2. Representations of integrated path during outgoing phase in EC
3. Representations of integrated path during incoming phase in EC
4. Representations of integrated path during outgoing phase in HC
5. Representations of integrated path during incoming phase in HC
6. Representations of goal proximity during outgoing phase in EC
7. Representations of goal proximity during outgoing phase in HC
8. Representations of goal proximity during incoming phase in HC
9. Landmark representations in RSC

We included (1) in the model as it has been suggested that the grid cell system is particularly involved in PI (Hafting *et al.*, 2005; Bush *et al.*, 2015). We included (3) in the model since it was significantly correlated with mean drop error across participants (*r*_34_ = -0.541, *P* = 0.001). Because (2), (5), and (7) were correlated with (1) (*r*_34_ = 0.50, *P* = 0.002), (1) (ρ_34_ = -0.38, *P* = 0.024), and (3), (*r*_34_ = -0.341, *P* = 0.045), respectively, they were excluded to avoid multicollinearity. Consequently, we started the model build-up using predictors (1), (3), (4), (6), (8), and (9). These predictors were not correlated (all *r*_34_ or ρ_34_ ≤ 0.32, all *P* ≥ 0.061). We allowed for interactions between the fMRI representations and incoming distance as well as subtask, but not for interactions between the fMRI representations. In the next step, we aimed at excluding predictors from the model that were not relevant for predicting path integration performance in order to build a parsimonious model (Bates *et al.*, 2015): We started selecting predictors by applying an arbitrary significance threshold of *α* = 0.1, which resulted in removing (4) and (6). We proceeded with the following procedure: We started with the highest order interaction term and removed it if it was not significant at *α* = 0.05. We calculated a new model and successively continued removing lower order terms (as long as they were not part of a higher order interaction) until there was no non-significant term that could be removed. This resulted in a final model including subtask, incoming distance and (1), (3), (8), (9) as well as their interactions with subtask and incoming distance as predictors (model 6).

### Creation of ROI masks and structural MRI analysis

Anatomical ROIs were created using a semiautomatic approach as implemented in Freesurfer (https://surfer.nmr.mgh.harvard.edu/). Briefly, structural MRIs were processed using the analysis pipeline of Freesurfer v6.0 that involves intensity normalization, registration to Talairach space, skull stripping, white matter (WM) segmentation, tesselation of the WM boundary, and automatic correction of topological defects (Fischl *et al.*, 2004). Pial surface misplacements and erroneous WM segmentation were manually corrected on a slice-by-slice basis to enhance the reliability of cortical thickness and hippocampal volume measurements. Segmented brain images were parcellated into cortical and subcortical regions (Desikan *et al.*, 2006; Destrieux *et al.*, 2010). This allowed us to obtain volume measurements of HC and EC. For the RSC ROI, we used the posterior-ventral part of the cingulate gyrus derived from the Destrieux atlas (following Shine *et al.*, 2016).

Human EC contains structurally and functionally distinct subparts (Maass *et al.*, 2015; Navarro Schröder *et al.*, 2015). Therefore, we subdivided the EC using a template of anterior-lateral EC (alEC) and posterior-medial EC (pmEC) based on functional connectivity with these subregions, which represent the human homologue of rodent lateral and medial EC (Navarro Schröder *et al.*, 2015), respectively. To obtain subject-specific masks, the template was mapped from Montreal Neurological Institute (MNI) space into individual subject’s native space. To ensure that only voxels located in gray matter were analyzed, the ROI masks were thresholded and intersected with a gray matter mask based on Freesurfer’s cortical parcellation. Each mask was manually inspected to ensure anatomical correctness.

To perform fMRI ROI analyses, ROIs were coregistered to the mean functional image of the respective participant so that all ROI analyses were performed in native space. For the sMRI analysis, we calculated relative gray matter volume by dividing the volume of the ROI by the whole brain volume of the respective participant (Freesurfer variable: “BrainSegVolNotVent”) as in previous studies (Kunz *et al.*, 2017).

### fMRI analysis

#### Preprocessing

Preprocessing of fMRI data was performed using SPM12 and included slice time correction and spatial realignment. For the whole-brain analysis, we normalized fMRI scans to MNI space using parameters from the normalization procedure of the segmented structural T1 image. Spatial smoothing with a 5 mm isotropic Gaussian kernel was applied to the normalized fMRI data and the fMRI data in native space. Normalized functional images were used to perform whole-brain analyses, while functional images remained in native space for the ROI analyses.

#### General linear models

Functional images were analyzed via two separate general linear models (GLMs).

##### PI model (Figure S6)

We used four different regressors to model the start phase, the outgoing phase, the incoming phase, and time periods of no movement, separately for each of the three subtasks and each of the 2 runs (24 regressors). Time periods of no movement were defined as those time periods when movement speed was at zero or minimally above zero (<1 percentile of the subject-specific movement speeds). In order to include parametric regressors for the moment-to-moment changes in integrated path and goal distance (see below), the regressors for outgoing phase and incoming phase had separate onsets at each movement time point (every 200 ms) and a duration of zero.

We hypothesized that regions involved in PI would represent the integrated path and/or the goal proximity of the current location. We further hypothesized that this representation should be more relevant, and thus more pronounced, when no supportive spatial cues were available (i.e., in the PPI subtask). In order to test this, we included one of two parametric modulators during the movement periods of the outgoing and the incoming phase: integrated path (i.e., the cumulative distance that has been traveled at each time point during the outgoing or incoming phase) or goal distance (i.e., the instantaneous Euclidean distance to the goal during the outgoing or incoming phase).

We assumed that goal proximity and integrated path were correlated, because this is often the case in the real world. To test this assumption empirically, we correlated goal proximity and integrated path across the entire time of the experiment within each participant. We tested each participant’s empirical Spearman ρ-value against a surrogate distribution of ρ-values established by shuffling the data time series for 10,000 times. We transformed the resulting *P*-value into a *z*-value. The mean empirical *z*-value across participants was tested against a surrogate distribution of mean *z*-values established by random sign flips for 10,000 times.

As expected, we found that goal proximity and integrated path were related in our task (ρ = 0.38, *Z* = 3.72, *P* < 0.001). Therefore, we built a third GLM that included both parametric modulators at the same time. Goal proximity was entered first, and integrated path was entered as a second, orthogonalized parametric modulator. We then correlated the β-values for the integrated path regressor from this GLM with the β-values from the GLM that only included the integrated path as parametric modulator to show that representations of integrated path were not a side-effect of goal-proximity representations.

##### Subtask model (Figure S7)

We modelled “start phase”, “outgoing phase”, “incoming phase”, and “feedback” separately for each of the three subtasks and each of the 2 runs (24 regressors). The duration of these regressors corresponded to the duration of the respective phases in each trial. Additionally, in the outgoing phase, we included “PI difficulty” as a parametric modulator, reflecting the number of distractor trees in each trial (with values of 1-5). This parametric modulator resembles the “outgoing distance” in the behavioral analyses and was included to reduce error variance in the GLM. We tested two contrasts: “BPI>PPI” and “LPI>PPI”.

All parametric modulators were normalized to values between 0 and 1 and mean-centered (Büchel *et al.*, 1998). All regressors were convolved with the haemodynamic response function before entering the GLM. As nuisance regressors, we included motion parameters as estimated in the realignment procedure and mean value of white matter and corticospinal fluid. We did not model temporal derivatives.

#### ROI analysis

For the ROI analysis, we extracted mean β-values (SPM con-files) from our three ROIs (EC, HC, RSC) in native space and compared them against zero across participants by means of *t*-tests for normally distributed data or Wilcoxon’s ranked sum tests otherwise. In order to compare mean β-values across subtasks, we used repeated measures analysis of variance (ANOVA) or Friedman ANOVA if data did not fulfil normality assumption. To test for differences between two conditions post-hoc, we used *t*-tests (or Wilcoxon ranked sum tests if normality assumption was violated).

Whenever we used the same model for several ROIs or several contrasts, we corrected for multiple comparisons using FDR (Benjamini and Hochberg, 1995; corrected for the number of contrasts and the number of ROIs, *α* < 0.05) as implemented in R and adjusted the *P*-values accordingly.

For the PI model, we corrected for 6 multiple comparisons when assessing the relationship between mean β-values and the parametric modulators (integrated path or goal proximity, respectively). These 6 comparisons correspond to 3 ROIs (EC, HC, RSC) and 2 contrasts against zero (one parametric modulator during the outgoing phase, one parametric modulator during the incoming phase).

Only if a contrast tested against zero showed a significant effect for an ROI (corrected for 6 multiple comparisons), we tested for differences between subtasks (one-way repeated measures ANOVA with subtask as within subjects factor), and again corrected for the number of ROIs. We performed two types of post-hoc comparisons: (1) We compared the subtasks against each other and corrected for 3 comparisons (PPI vs. BPI, PPI vs. LPI, BPI vs. LPI); (2) we tested every subtask against zero and likewise corrected for 3 comparisons (PPI vs. 0, BPI vs. 0, LPI vs. 0).

For the subtask model, we assessed the influence of either a boundary or a landmark in the environment on BOLD activity and thus corrected for 6 multiple comparisons (3 ROIs: EC, HC, RSC; 2 contrasts: BPI>PPI, LPI>PPI). We did not directly compare the boundary and the landmark condition.

#### Whole brain analysis

In the subtask model, we performed whole brain analyses on the contrasts between subtasks. Contrast images from the first-level analysis of each participant were entered into a GLM, treating “subjects” as a random effect. Statistical parametric maps were initially thresholded at an FWE-corrected alpha level of *P* < 0.05 across the whole brain. We considered clusters significant at *P* < 0.05, FWE-corrected (extent threshold of 5 voxels). For all significant clusters, we provide maximum probability tissue labels with MNI coordinates derived from the Neuromorphometrics atlas as implemented in SPM12 (http://www.oasis-brains.org/; http://Neuromorphometrics.com/). Given our strong hypothesis that EC, HC, and RSC would be involved in our task, we used small volume correction with a mask of the EC, HC, and RSC. We derived these masks from Freesurfer by warping subject-specific ROIs to standard space, averaging the mask across subjects, and thresholding the composite mask, such that a selected voxel was included in the masks of at least 50% of the participants. Thereby, we ensured that results of the ROI analysis and of the whole brain analysis would correspond as closely as possible.

#### Grid-like representations

In order to detect GLRs in the fMRI data, we performed a representational similarity analysis [RSA (Kriegeskorte and Kievit, 2013; Haxby, Connolly and Guntupalli, 2014)] following a previous study (Bellmund *et al.*, 2016). The rationale underlying this analysis is that the activity of a grid cell population should be similar during movements that are *n*\*60° offset from each other (due to the six-fold rotational symmetry of the grid pattern), where *n* = {0, 1, …, 6}. In other words, the activity of a grid cell population should be relatively similar at angular steps of 60°. By contrast, the activity of a grid cell population should be relatively dissimilar during movements that are *n*\*60°+30° offset from each other. In the following, the former condition is termed “mod(α,60°)=0°” and the latter condition is termed “mod(α,60°)=30°” (where α refers to the angular difference between movement directions, and “mod” indicates the modulo operator; **Figure 5A**).

Analysis steps comprised (1) extracting the voxelwise signal within the ROIs for every fMRI volume; (2) calculating mean orientation, speed, and trial number associated with each fMRI volume; (3) excluding volumes associated with slow movement (< 33% average movement speed); (4) creating means of volumes within bins of 5° orientation based on the information about mean orientation; (5) calculating the angular difference of movement directions between volume bins; (6) calculating Fisher-*z*-transformed Pearson correlations between the volume bins; and (7) deleting correlations of the same trial in order to reduce effects of temporal autocorrelations between subsequent fMRI volumes. We compared pattern similarities of angular differences of ±15° from the mod(α,60°)=0° condition, for which we expected higher pattern similarity, against angular differences of ±15° from the mod(α,60°)=30° condition, for which we expected lower pattern similarity.

As a control, we re-performed the analysis after excluding correlations based on angular differences of ±15° from 0° in order to exclude the possibility that GLRs were driven by similarities of activity during movements in the same direction (in 360° space), possibly reflecting a head direction signal. To examine the specificity of six-fold rotational symmetry, we performed further control analyses that tested for other types of rotational symmetry (4-fold, 5-fold, 7-fold, and 8-fold rotational symmetry).

In order to exclude possible effects of autocorrelations as a confound, we tested whether the temporal proximity between fMRI volumes assigned to the two conditions was similar. We thus calculated the mean TR difference between every possible combination of angular differences and averaged within the mod(α,60°)=0° condition and the mod(α,60°)=30° condition for every participant. We calculated a paired *t*-test across participants to ensure that the temporal proximity did not differ between the two conditions. Finally, for each participant separately, we checked for uniform sampling of movement directions in 360° and in 60° space. To this end, we transformed the movement directions (in 360° space or in 60° space) into bins of 5° and tested if the orientation bins deviated from uniformity by means of a Rayleigh test. This resulted in an empirical *z*-value that we tested against a surrogate distribution established by shuffling the orientation bins 10,000 times. Comparing the empirical *z*-value against the distribution of surrogate *z*-values resulted in one *P*-value for each participant reflecting how normal or extreme the empirical *z*-value of each participant was.

We calculated spatial and temporal signal-to-noise ratios (SNRs; following Bellmund *et al.*, 2016) by extracting the voxelwise signal of alEC and pmEC for each hemisphere and either calculating the mean and the s.d. across timepoints (temporal SNR) or across voxels (spatial SNR). We compared SNRs between EC subregions (alEC vs. pmEC) and between hemispheres (left vs. right) using a two-way ANOVA. By means of a Pearson correlation, we tested whether the strength of GLRs was significantly related to the SNRs of pmEC.

